# Mathematical modelling of COVID-19 with periodic transmission: The case of South Africa

**DOI:** 10.1101/2022.06.22.22276298

**Authors:** Belthasara Assan, Farai Nyabadza

## Abstract

The data on SARS-CoV-2 (COVID-19) in South Africa shows seasonal transmission patterns to date, with the peaks having occurred in winter and summer since the out-breaks began. The transmission dynamics have mainly been driven by variations in environmental factors and virus evolution, and the two are at the center of driving the different waves of the disease. It is thus important to understand the role of seasonality in the transmission dynamics of COVID-19. In this paper a compartmental model with a time dependent transmission rate is formulated and the stabilities of the steady states analysed. We note that if *R*_0_ < 1, the disease-free equilibrium is globally asymptotically stable, and the disease completely dies out and when *R*_0_ > 1, the system admits a positive periodic solution, and the disease is uniformly or periodically persistent. The model is fitted to data on new cases in South Africa for the first four waves. The model results clearly indicate the need to consider seasonality in the transmission dynamics of COVID-19 and its importance in modelling fluctuations in the data for new cases. The potential impact of seasonality in the transmission patterns of COVID-19 and the public health implications are discussed.

## 1. Introduction

COVID-19 is an infectious disease caused by severe acute respiratory syndrome coronavirus 2 (SARS-CoV-2). The disease was first identified in December 2019 in Wuhan, the capital of Hubei, China, and has then spread globally, resulting in the ongoing 2020 pandemic outbreak [1]. It has caused substantial mortality and a major strain on healthcare systems [2, 3]. At the opposite extreme, many countries lack the testing and public health resources to mount similar responses to the COVID-19 pandemic, which could result in unhindered spread and catastrophic outbreaks. The disease has spread globally and the virus has affected over 500 million people and caused the death of more than 6 million people as of April, 2022.

The only pharmaceutical interventions available are vaccination and some anti-viral medicines [4]. Non-pharmaceutical interventions such as social distancing, intensive testing and isolation of cases [8] have been the mainstay in the control and management of COVID-19 in many African countries. The COVID-19 vaccines primarily prevents development of severe disease that leads to hospitalisation but not necessarily the infection. That means, vaccines does not block people from transmitting the pathogen to others. As a result, social distancing has quickly become an important consideration in mathematical modelling [5, 6, 7]. Social distancing is maintaining a physical distance between people and reducing the number of times people come into close contact with each other. It usually involves keeping a certain distance from others (the distance specified differs from country to country and can change with time) and avoiding gathering together in large groups [11]. This minimises the probability that an uninfected person will come into physical contact with an infected person, thus suppressing the disease transmission, resulting in fewer hospitalisation and deaths.

Studies done on social distancing shows that the adoption of relaxed social distancing measures reduces the number of infected cases but does not shorten the duration of the epidemic waves. Increasing social distancing could reduce the number COVID-19 new cases by 30% [17]. Researchers believe that social distancing offers more advantages than drawbacks, since it can serve as a non-pharmacological tool to reduce the disease’s proliferation rate.

Data on COVID-19 cases has shown trends of seasonal variations. Environmental factors and other climatic factors, are often seasonal and could significantly affect disease dynamics [15, 16]. The seasonal cycles are an ubiquitous feature of influenza and other respiratory viral infections, particularly in temperate climates [17]. Like any other respiratory viral infection, COVID-19 has been found to exhibit some form of seasonality [18, 19]. The outcome of the research in [18, 19] suggest that transmissibility of the COVID-19 can be affected by several meteorological factors, such as temperature and humidity. These conditions are probably favourable for the survival of the virus in the transmission routes. Research done by [20] found out that 60% of the confirmed cases of COVID-19 occurred in places where the air temperature ranged from 5°*C* to 15°*C*, with a peak in cases at 11.54°*C*. Moreover, approximately 73.8% of the confirmed cases were concentrated in regions with absolute humidity of 3*g*/*m*^3^ to 10*g*/*m*^3^. SARS-CoV-2 however, appears to be spreading toward higher latitudes. In this paper, we consider South Africa, with two main climate seasons in a year. And during these periods there has always been a surge of infections.

Seasonal patterns expose the limitations of many recent COVID-19 models that do not incorporate seasonality. We therefore propose a COVID-19 model with periodic transmission, that incorporates periodicity in the disease transmission pathway. In this case, the incidence is subject to periodicity. We analyze the basic reproduction number, *R*_0_, for the model and establish that *R*_0_ is a sharp threshold for COVID-19 model with periodic transmission. The method of analysis for extinction and persistence results for periodic epidemic systems is inspired by the research done in [21, 22].

This paper is organized as follows. In Section 2, we propose a new model for COVID-19 with periodic social distance parameter in the force of infection. A qualitative analysis of the model is investigated in Section 3. The usefulness of our model is then illustrated in Section 4 where we fit our model to the incidence data from South Africa. We conclude the paper in Section 5.

## 2. The Mathematical model

### 2.1. Model formulation

We consider the total human population at time *t* defined by *N*(*t*). The total population is divided into five compartments of: the susceptibles, *S* (*t*), the exposed, *E*(*t*), the asymptomatic infectives *I*_*a*_(*t*), the symptomatic infectives *I*_*s*_(*t*) and those who would have recovered from the infection, *R*(*t*), so that

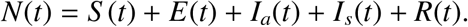

The following compartmental diagram shows the flow of populations between compartments.

Table 1 shows the parameter used in formulating the model.

**Table 1.** Description of parameters used in the model

| Parameter | Description |
| --- | --- |
| $\Lambda$ | Recruitment rate of humans. |
| $\mu$ | Natural mortality rate for humans. |
| $\delta(t, I_a, I_s)$ | The disease transmission rate for humans dependent on time. |
| $\kappa$ | Progression rate of exposed individuals to the asymptomatic and symptomatic classes. |
| $\gamma_a, \gamma_s$ | rate of recovery for asymptomatic and symptomatic individuals respectively. |
| $\alpha$ | Rate of transfer from asymptomatic to symptomatic individuals. |
| $\psi$ | Disease induce death rate. |
| $m(t)$ | The periodic rate at which individuals distance themselves from each other. |
| $\eta$ | Relative infectivity parameter for the symptomatic compared to the asymptomatic. |

From the compartmental model in Figure 1 the following non-autonomous dynamical system is derived to describe the dynamics of the transmission of COVID-19 with periodic social distancing in the force of infection:

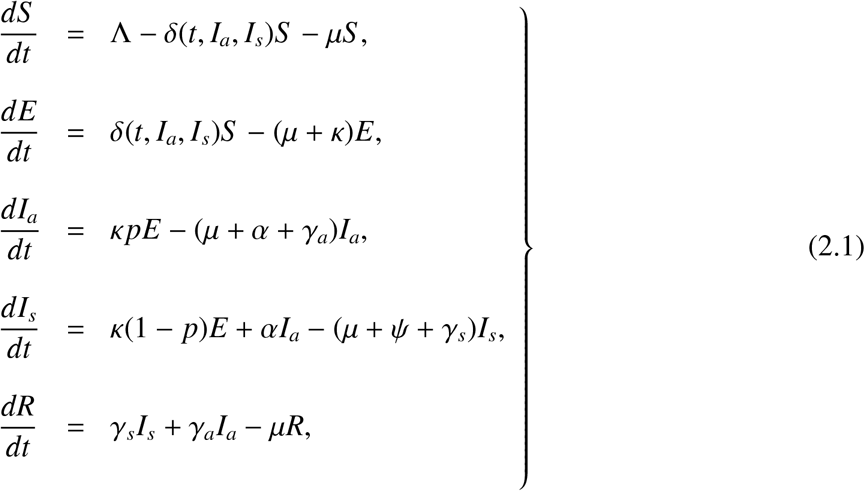

where

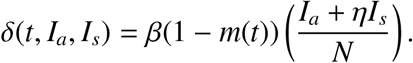

**Figure 1.**
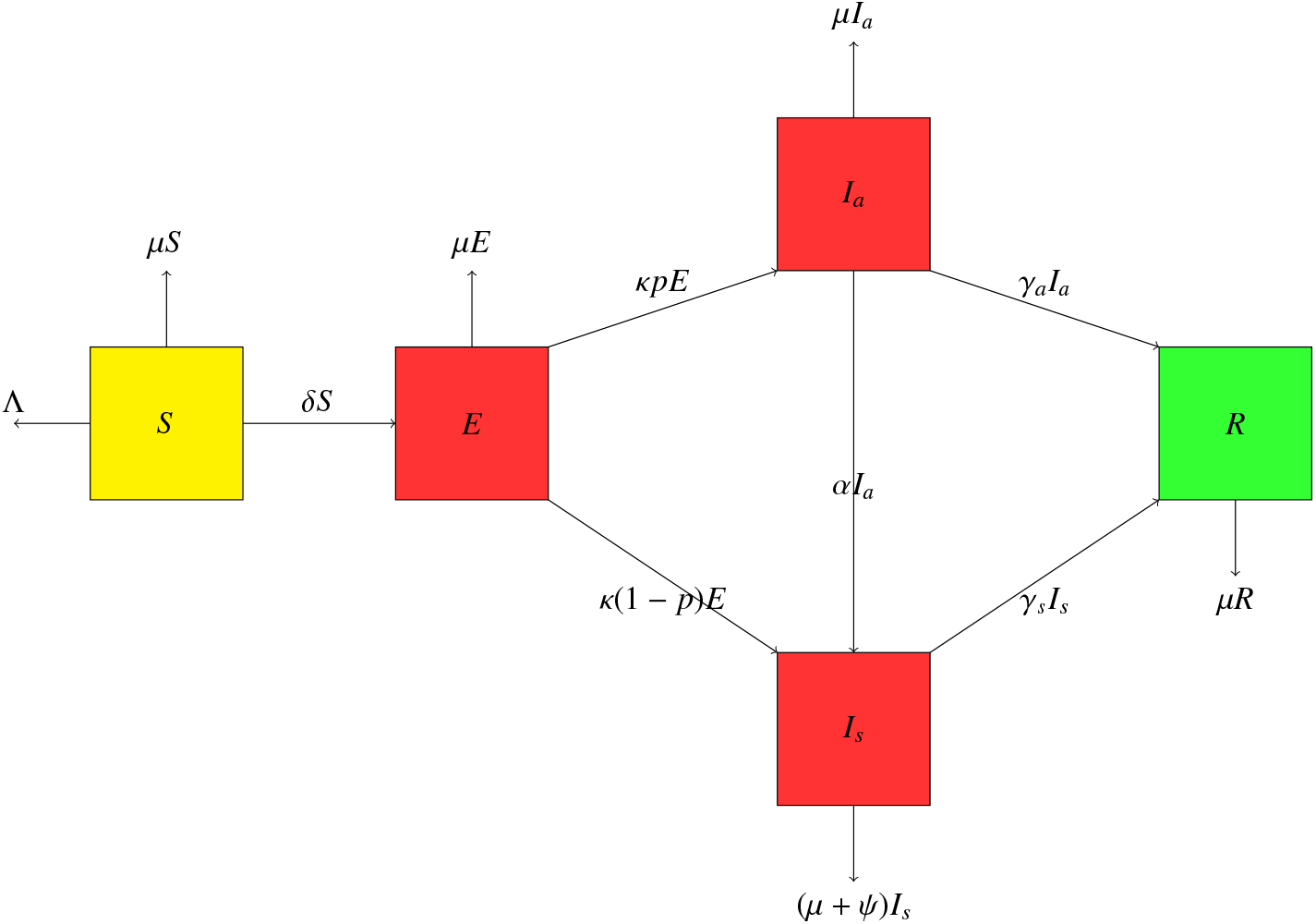
Compartmental model of COVID-19 with periodic transmission

The incidence function *δ*(*t, I*_*a*_, *I*_*s*_) determines the rate at which new cases of COVID-19 are generated. The parameter *η* measures the difference in infectivity between *I*_*a*_ and *I*_*s*_. The rates *m*(*t*) is a periodic function of time with a common period, 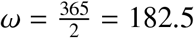 days. Periodic transmission is often assumed to be sinusoidal, such that

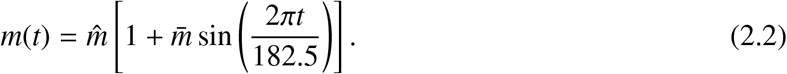

The product 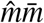, is the amplitude of the periodic oscillations in *m*(*t*). There is no periodic infections when 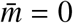. Here 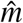 is thus the baseline value or the time-average value of social distancing.

## 3. Assumptions, equilibrium points and analysis

The function *δ*(*t, I*_*a*_, *I*_*s*_) is differential and periodic in time with period *ω*. That is,

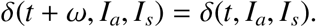

To make biological sense, we assume that the function *δ* satisfy the following three conditions (*A*1 − *A*3) for all *t* ≥ 0:

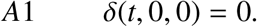

Setting all the derivatives of dynamical system to zeros and setting all infected class to be zero that is, (*E, I*_*a*_, *I*_*s*_) = (0, 0, 0), gives the disease free equilibrium

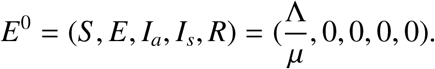

Assumption (*A*1), ensures that the model has a unique and constant *E*^0^.

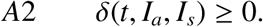

The assumption *A*2 ensures a non-negative force of infection. So 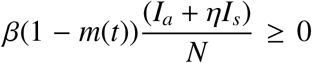 with the condition that 0 ≤ *m*(*t*) ≤ 1, ∀*t* > 0.

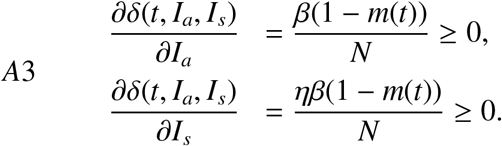

Assumption *A*3 states that the rate of new infection increases with both the infected population size. Assumption *A*4, below shows that geometrically, the surface that represents the force of infection, *δ*, lies below its associated tangent plane at the origin. This means that the remainder term, *R*_1_, from the truncated Taylor expansion of *δ* when the degree is equals to one is non-positive. The second partial derivatives of the force of infection are

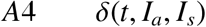

is concave for any *t* ≥ 0 ; i.e. the matrix

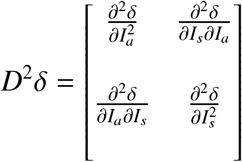

is negative semi-definite everywhere. Since

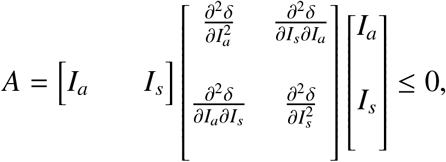

we have,

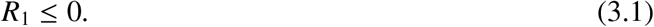

It follows that if *R*_1_ ≤ 0, the matrix *A* is negative semi-definite. The model shows that a single infected individual is sufficient for a positive infection rate.

### 3.1. The basic reproduction number, using the next infection operator, L

Wang and Zhao [9] extended the framework in [10] to consider an epidemiological model in periodic environments. Let Φ_−*V*(*t*)_ and *ρ*(Φ_−*V*(*ω*)_) be the monodromy matrix of the linear *ω*−periodic system 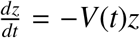 and spectral radius of Φ_−*V*(*ω*)_ respectively. Assume *Y*(*t, s*), *t* ≥ *s*, is the evolution operator of the linear *ω*−periodic system

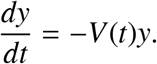

That is, for each *s* ∈ ℝ, the 3 × 3 matrix *Y*(*t, s*) satisfies,

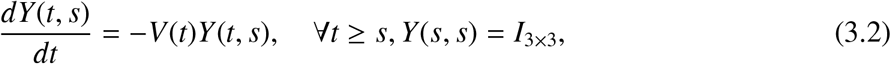

where *I*_3×3_ is identity matrix. In view of the fluctuating environments, we assume that Φ(*s*) is the initial distribution of infectious individuals and is *ω*− periodic in *s*. Then *F*(*s*)Φ(*s*) is the rate of new infections produced by the infected individuals who were introduced at time *s*. Given that *t* ≥ *s*, then *Y*(*t, s*)*F*(*s*)Φ(*s*) gives the distribution of those infected individuals who were newly-infected at time *s* and remain in the infected compartments at time *t*. It follows that:

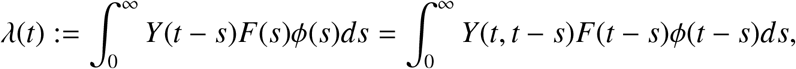

is the cumulative number of new infections at time *t* produced by all those infected individuals *ϕ*(*s*) introduced at time previous to *t*. Let *C*_*ω*_ be the ordered Banach space of all *ω*−periodic functions from ℝ to ℝ^3^, which is equipped with the maximum norm ||. || and the positive cone

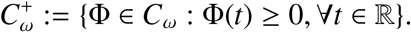

Then we can define a linear operator *L* : *C*_*ω*_ → *C*_*ω*_ by

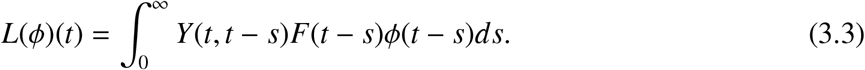

Here, *L* is the next infection operator, and we define the basic reproduction number as

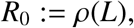

the spectral radius of *L*. We now define *F*(*t*) and *V*(*t*) according to equation (2.1).

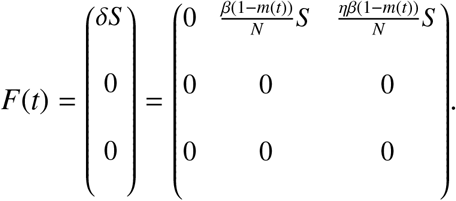

Substituting 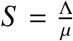, we get:

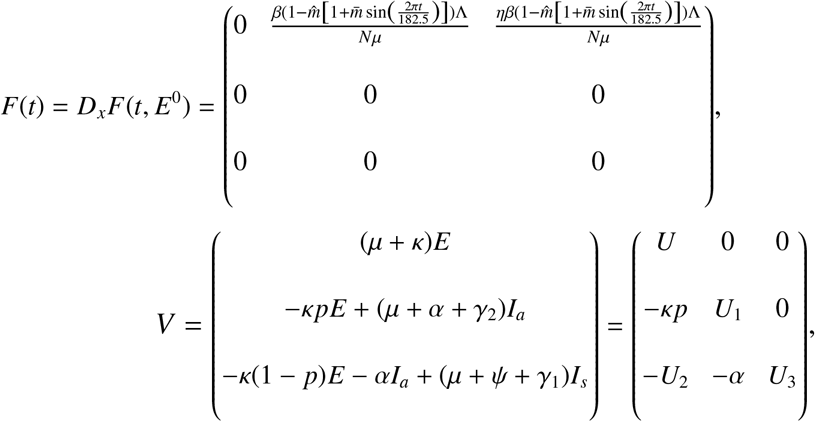

where

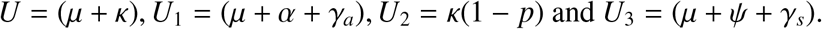

Also,

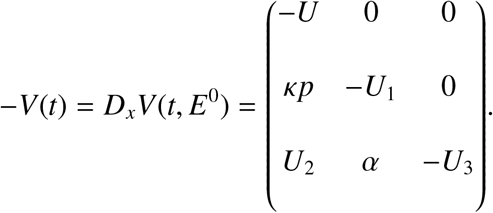

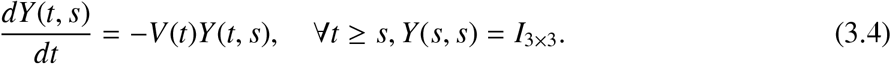

Solving the ODE with it’s initial conditions we have,

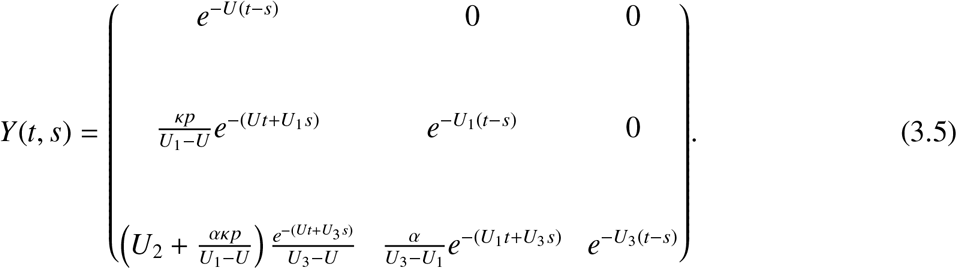

We numerically evaluate the next infection operator by

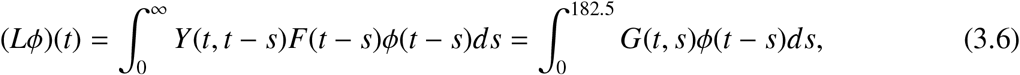

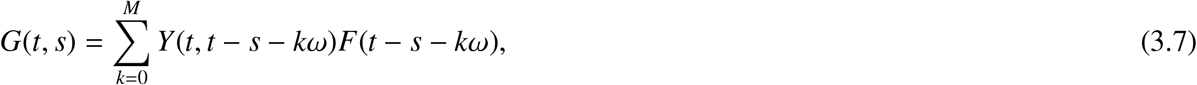

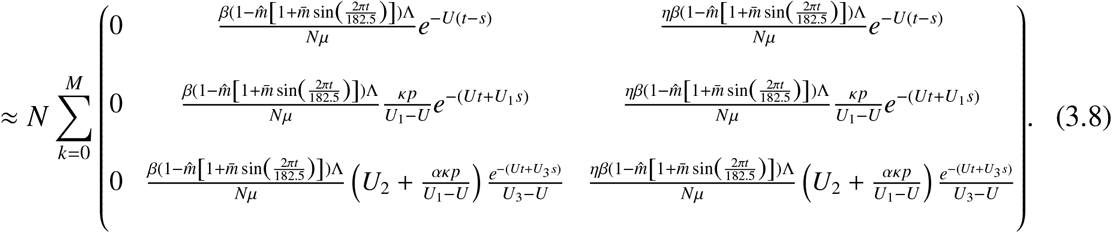

For some positive integer *M*

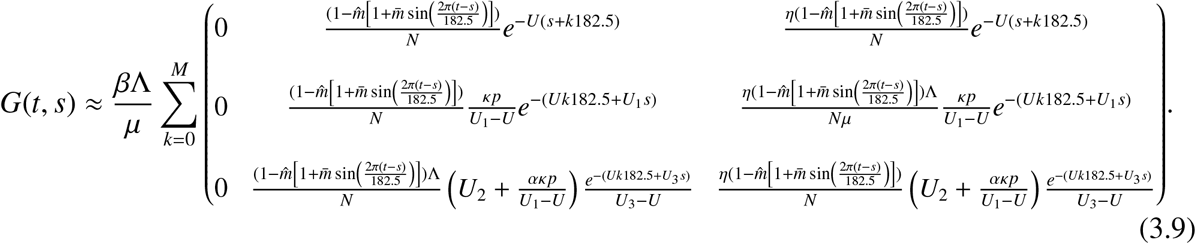

To compute the basic reproduction number *R*_0_, we reduce the operator eigenvalue problem to a matrix eigenvalue problem in the form of *Ax* = *λx*, where matrix *A* can be constructed by arranging the entries of the function *G*. The basic reproduction number *R*_0_ can then be approximated by numerically calculating the spectral radius of the matrix *A*. The monodromy matrix of differential equation (3.2) is

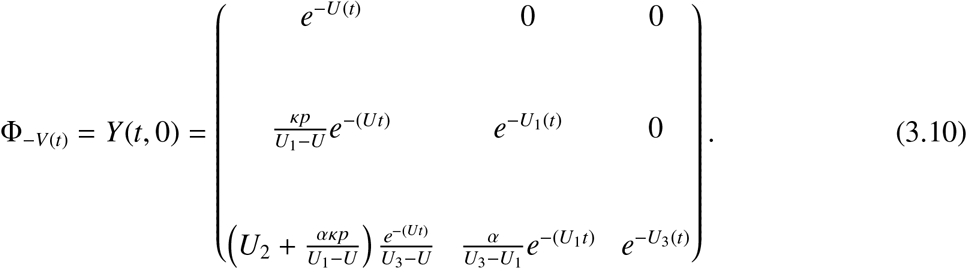

It follows that the eigenvalues of any lower triangular matrix are the diagonal elements. Thus, the spectral radius of 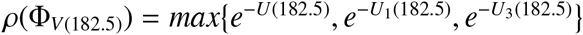

#### Lemma 1.

*The following statements are valid for Equation (2.1)*

*(i) R*_0_ = 1 *if and only if ρ*(Φ_*F*−*V*_ (*ω*)) = 1.

*(ii) R*_0_ > 1 *if and only if ρ*(Φ_*F*−*V*_ (*ω*)) > 1.

*(iii) R*_0_ < 1 *if and only if ρ*(Φ_*F*−*V*_ (*ω*)) < 1.

*(iv) The disease free equilibrium, E*^0^, *is locally asymptotically stable if R*_0_ < 1, *and unstable if R*_0_ > 1.

*Proof*. Since Model 2.1 satisfies conditions (A1)—(A7) in [25], it follows that Theorem 2.2 of [9], hold for all conditions D

### 3.2. Disease extinction

In this subsection, we analyse the global stability of the disease free equilibrium point for the system (2.1). This gives condition for disease extinction. Suppose we have the matrix function

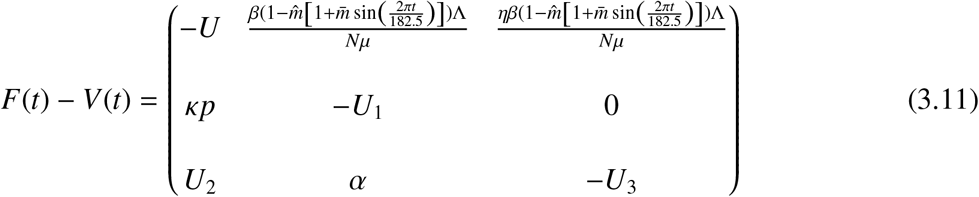

where *F*(*t*) = *Dℱ* (*E*^0^) and *V*(*t*) = *D𝓋*(*E*^0^). The disease free equilibrium *E*^0^ is globally asymptotically stable if all the eigenvalues of the matrix *DE*^0^ = {*F*(*t*)−*V*(*t*)} have positive real parts. Clearly, the matrix function of Equation (3.11) is irreducible, continuous, cooperative, and ω-periodic. Let Φ_(*F*−*V*)_(*t*) be the fundamental solution matrix of the linear ordinary differential system:

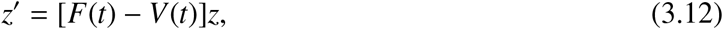

where *𝓏* = [*E, I*_*a*_, *I*_*s*_]^*T*^ and *ρ*(Φ_*F*−*V*_ (*ω*)) is the spectral radius of (Φ_*F*−*V*_ (*ω*)).

#### Lemma 2.

*Let* 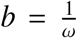 In *ρ*(Φ_*F*−*V*_ (*ω*). *Then there exist a positive ω*−*periodic function v*(*t*) *such that e*^*bt*^*v*(*t*) *is a solution to Equation (3.12)*.

*Proof*.

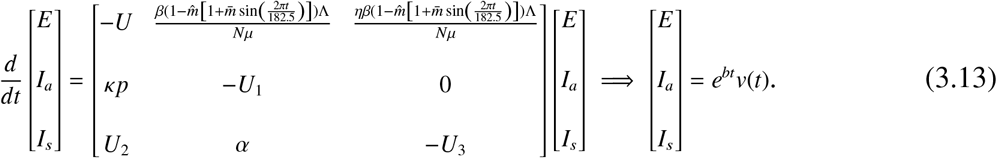

Since *R*_0_ < 1 then, *ρ*(Φ _*f*_ (182.5)) < 1, then *b* < 0, it follows that *R*_0_ < 1 implies

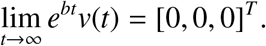

Considering and using *A*4, *R*_1_ ≤ 0, this the follows:

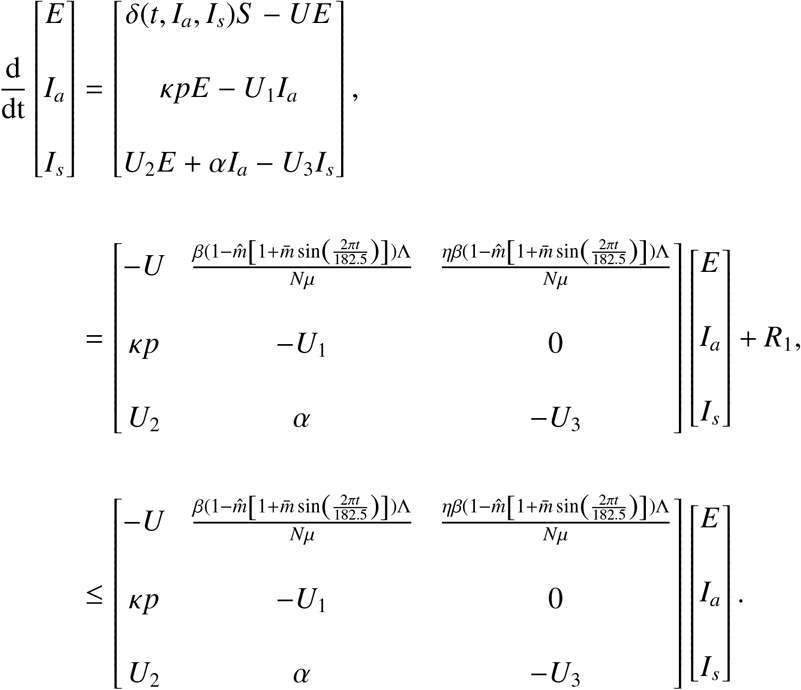

From [Theorem 2.2 [9]], *R*_0_ < 1 if and only if *ρ*(Φ_(*F*−*V*)_(182.5)) < 1. Therefore, *b* < 0. It is clear that

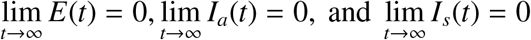

Next, we consider the last equation in Equation (2.1). For any *ε* > 0, there exists *T* > 0 such whenever *t* > *T*, we have

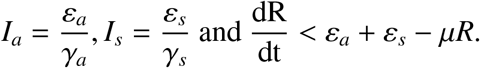

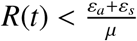 for *t* > *T*. Since *ε* > 0 is arbitrary so

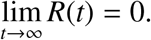

Since the total population *N*(*t*) = *S* (*t*) + *E*(*t*) + *I*_*a*_(*t*) + *I*_*s*_(*t*) + *R*(*t*), we have that

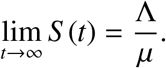

We thus have the following result.

#### Theorem 1.

*If R*_0_ < 1, *then the disease-free equilibrium of Equation of 2.1 is globally asymptotically stable*.

Theorem 1 shows that the disease will completely die out as long as *R*_0_ < 1. This further implies that reducing and keeping *R*_0_ below 1 would be sufficient to eradicate COVID-19 infection even with a periodic transmission.

### 3.3. Disease persistence

In this subsection, we consider the dynamics of the Equation (2.1) when *R*_0_ > 1, where *E*^1^ be the endemic equilibrium point for Equation (2.1). Let us consider also the following set:

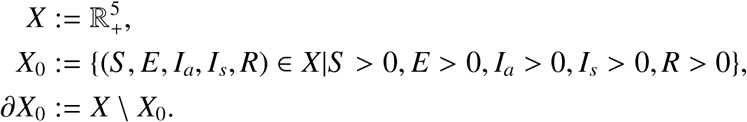

Let *u*(*t, ϕ*) be the unique solution of Equation (2.1) with initial condition *ϕ*, Φ(*t*) semi-flow generated by periodic Equation (2.1) and *P* : *X* → *X* the Poincaré map associated with Equation (2.1), namely:

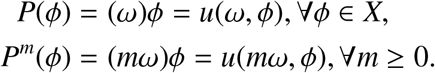

#### Proposition 3.4.

*The set X*_0_ *and* ∂*X*_0_ *are positively invariant under the flow induced by Equation (2.1)*.

*Proof*. For any initial condition *ψ* ∈ *X*_0_, by solving Equation (2.1), we derive that:

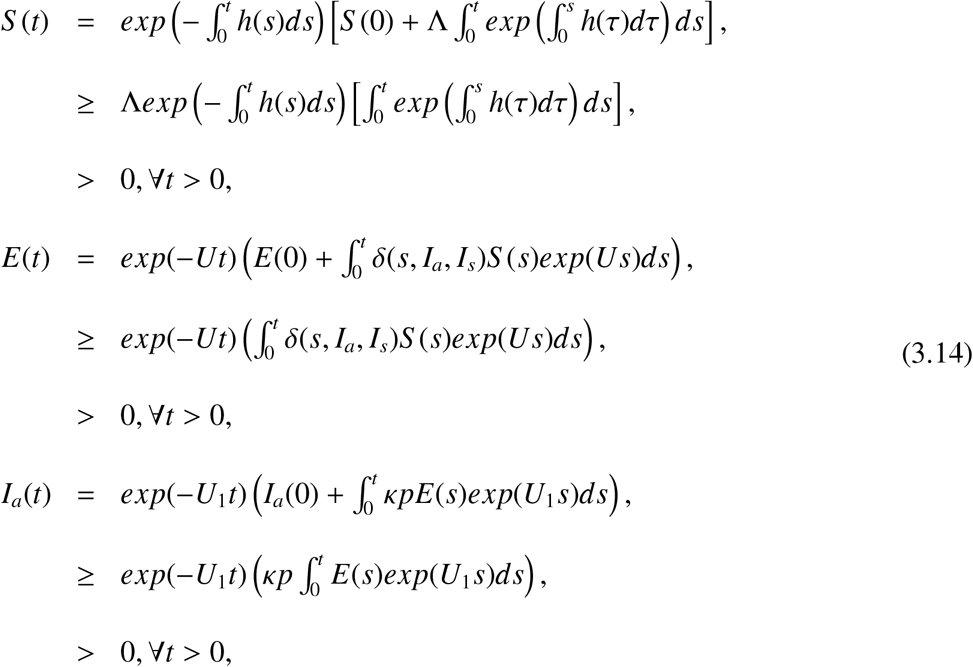

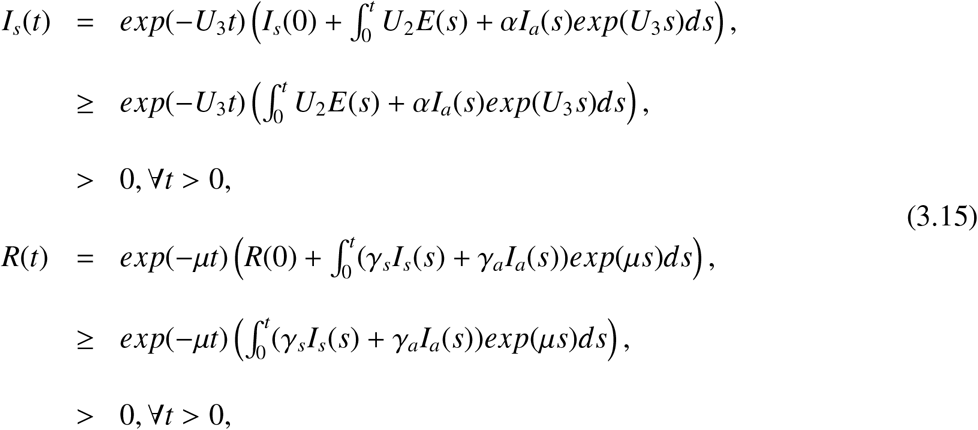

where, *h* = (*δ*(*t, I*_*a*_, *I*_*s*_) − *µ*)*S*.

Thus *X*_0_ is positively invariant. Since *X* is positively invariant and ∂*X*_0_ is relatively closed in *X*, it yields that ∂*X*_0_ is positively invariant.

The compact Ω, is defined as a positive invariant set for the Equation (2.1), which attracts all positive orbits in 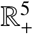 and the solutions are bounded. Ω attracts all positive orbits in *X* which implies that the discrete-time system *P* : *X* → *X* is point dissipative. Moreover, 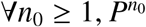 is compact, it then follows that *P* admits a global attractor in *X*.

#### Lemma 3.

*If R*_0_ > 1, *there exists η* > 0 *such that when* | |*ϕ* − *E*^1^| | ≤ *η*, ∀*ϕ* ∈ *X*_0_ *we have*

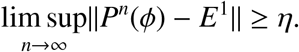

*Proof*. Suppose by contradiction that lim sup||*P*^*n*^(*ϕ*) − *E*^1^|| < *η* for some *ϕ* ∈ *X*_0_. Then, there exists an integer *N*_1_ ≥ 1 such that for all 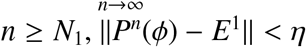. By the continuity of the solution *u*(*t, ϕ*), we have ||*u*(*t, P*^*n*^(*ϕ*)) − *u*(*t, E*^1^) ≤ *σ* ||for all *t* ≥ 0 and *σ* > 0. For all *t* ≥ 0, let *t* = *nω* + *t*′, where *t*′ ∈ [0, *ω*] and *n* = [*t*/*ω*]. [*t*/*ω*] is the greatest integer less or equal to *t*/*ω*. If ||*ϕ* − *E*^1^|| ≤ *η*, then by the continuity of the solution *u*(*t, ϕ*), we have

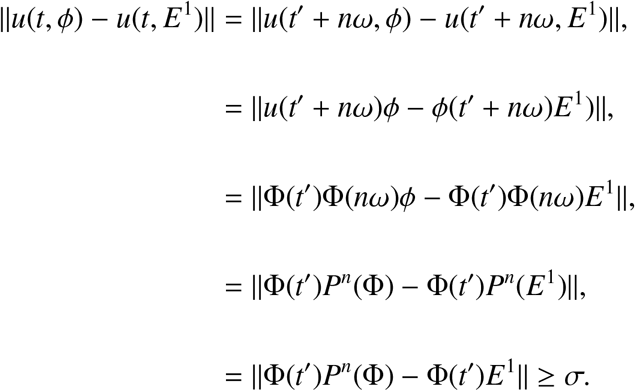

Moreover, there exists *σ*^∗^ > 0 such that for all *t* ∈ [0, *ω*],

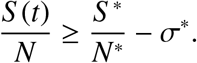

Hence, we obtain the following equation:

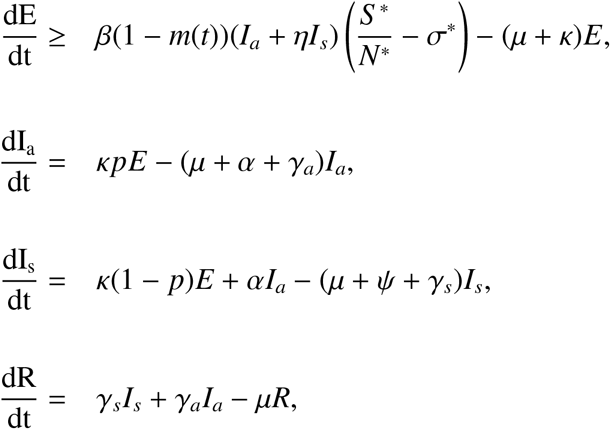

Let us consider the following auxiliary linear equation:

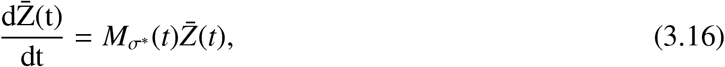

where

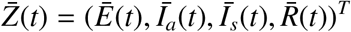

and 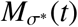 a matrix defined by

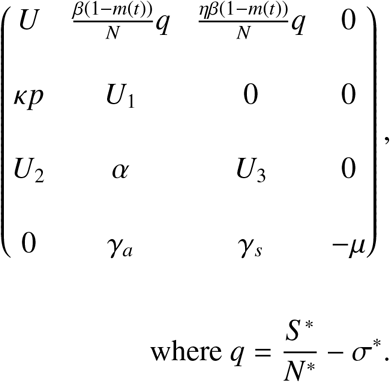

By Lemma 2, there exists a positive *ω*−periodic function *v*(*t*) such that 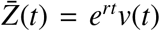 is a solution of Equation (3.16) with 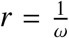 In *ρ*(Φ_*Mσ\**_ (*ω*)).*ρ*(Φ_*Mσ\**_ (*ω*))> 1 implies that *r* > 0. In this case, 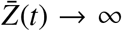 as *t* → ∞. Applying the theorem of comparison [23], we have

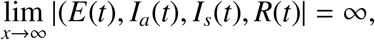

that contradicts the fact that solutions are bounded.

#### Theorem 2.

*If R*_0_ > 1, *there exists ξ* > 0 *such that any solution u*(*t, ϕ*) *with the initial condition ϕ* ∈ *X*_0_ *satisfies*

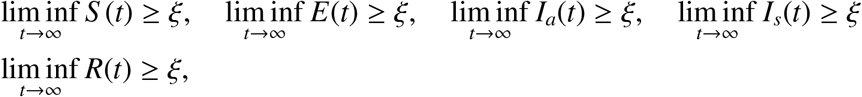

and the Equation (2.1) has at least one positive periodic solution.

*Proof*. Let us consider the following sets:

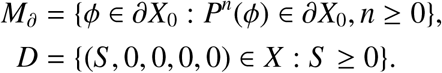

It is clear that *D* ⊂ *M*_∂_. So we must prove that *M*_∂_ ⊂ *D*. That means, for any initial condition *ϕ* ∈ ∂*X*_0_, *E*(*nω*) = 0 or *I*_*a*_(*nω*) = 0 or *I*_*s*_(*nω*) = 0 or *R*(*nω*) = 0 or, for all *n* ≥ 0. Let *ϕ* ∈ ∂*X*_0_,. Suppose by contradiction that there exists an integer *n*_1_ ≥ 0 such that *E*(*n*_1_*ω*) > 0 or *I*_*a*_(*n*_1_*ω*) > 0 or *I*_*s*_(*n*_1_*ω*) > 0 or *R*(*n*_1_*ω*) > 0, Then, by replacing the initial time *t* = 0 by *t* = *n*_1_*ω* in Equation (3.15) - (3.16), we obtain *S* (*t*) > 0, *E*(*t*) > 0, *I*_*a*_(*t*) > 0, *I*_*s*_(*t*) > 0, *R*(*t*) > 0 that contradicts the fact that ∂*X*_0_ is positively invariant. The equality *M*_∂_ = *D* implies that *E*^1^ is a fixed point of *P* and acyclic in *M*_∂_, every solution in *M*_∂_ approaches to *E*^1^. Moreover, Lemma 3 implies that *E*^1^ is an isolated invariant set in *X* and *W*_*s*_(*E*^1^) ∩ *X*_0_ = ∅. By the acyclicity theorem on uniform persistence for maps [[24], Theorem 3.1.1], it then follows that *P* is uniformly persistent with respect to (*X*_0_, ∂*X*_0_). So the periodic semiflow Φ(*t*) is also uniformly persistent. Hence, there exists *ξ* > 0 such that

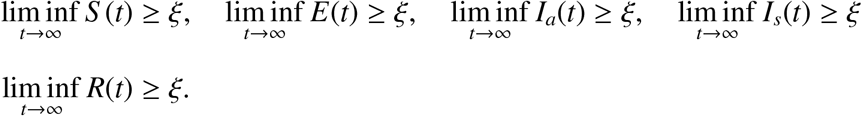

Furthermore, from [[24], Theorem 1.3.6], the system (2.1) has at least one periodic solution *u*(*t, ϕ*^∗^) with *ϕ*^∗^ ∈ *X*_0_. Now, let us prove that *S* ^∗^(0) is positive. If *S* ^∗^(0) = 0, then we obtain that *S* ^∗^(0) > 0 for all *t* > 0. But using the periodicity of solution, we have *S* (0) = *S* (*nω*) = 0 that is also a contradiction.

## 4. Numerical simulations

We now apply our model to the COVID-19 pandemic in South Africa. We use the outbreak data published daily by the National Institute for Communicable Diseases (NICD) [26]. These data sets contain the daily reported new cases, cumulative cases, and disease induced deaths for South Africa, for each province in South Africa. We shall however, in this project, consider the national data.

The numerical simulations were done using Python software. We use the force of infection with interventions, given by

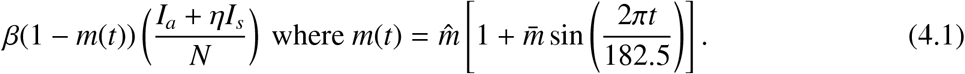

In our case where the intervention is social distancing, *m*, it is assumed to be periodic, due to the fluctuations in the existing data. The parameter *β*, is positive and denotes the maximum value of he transmission rate. We conduct numerical simulations for an epidemic period starting from 27^*th*^ March, 2020, when South Africa was locked and social distancing was observed in every place to 19^*th*^ April, 2022. Our time was set in days and given our period-*ω* to be 182.5 days, due to numbers of waves in a year. We fit the constructed model to a new cases of South Africa [26] in order to illustrate our mathematical results. The total population of South Africa in the year 2020 was 59.31 million [29]. The mortality rate *µ*, was obtained by using the life expectancy of South Africa which was found to be 64.3 years. The transmission rate was obtained through data fitting and all other parameters that were used in this section. The social distancing parameter *m*(*t*) and the proportion of exposed individuals *p* are all in the interval (0, 1).

To estimate the values for the rest of parameters not given above, we fit our model to the new cases reported daily in South Africa from 27^*th*^ March, 2020 to 19^*th*^ April, 2022. Studies reveals that, the true number of COVID-19 infections in Africa could be 97% higher than the number of confirmed reported cases [30, 31]. The initial conditions of all state variables are taken from the data. We set our initial conditions for each wave as:

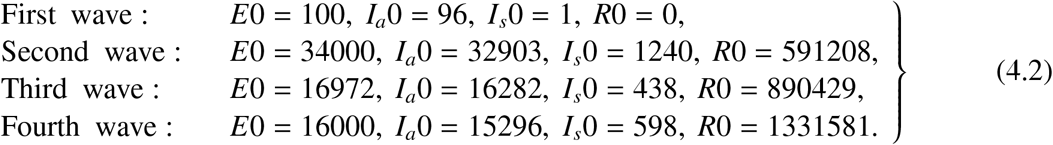

Initial conditions for susceptible and the asymptotic infectives are given by *S* 0 = *N*−(*E*0+*I*_*a*_0+*I*_*s*_0+*R*0) and 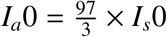 respectively.

Table 2 gives the start dates, end dates and the number of days in which each wave occur. We thus consider the system

**Table 2.** Time and days for the waves used in the estimation of initial conditions

| Waves | Start date | End date | Days |
| --- | --- | --- | --- |
| 1 | 27<sup>th</sup> March, 2020 | 11<sup>th</sup> September, 2020 | 178 |
| 2 | 11<sup>th</sup> September, 2020 | 10<sup>th</sup> April, 2021 | 200 |
| 3 | 11<sup>th</sup> April, 2021 | 29<sup>th</sup> October, 2021 | 200 |
| 4 | 30<sup>th</sup> October, 2021 | 19<sup>th</sup> April, 2022 | 171 |

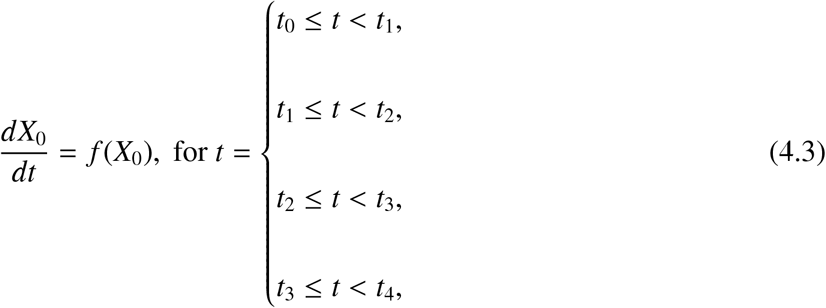

where

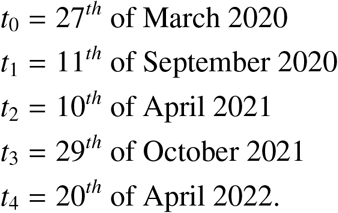

In Table 3, we discuss in detailed the values of the parameters used in Equation (2.1). During fitting of the mathematical model to the data, we divided *days* according to the number of waves we have. The table compares the transmission rate *β*, finding the first wave to have high transmission rate among wave 2 and 3, this is due to some preventive measures not implemented. In the presence of vaccination, social distance, and other preventive measures the fourth wave still tends to have a higher transmission rate compared to all the waves.

**Table 3.** Parameters Estimation of COVID-19 in South Africa for the first four waves in days

| Estimated parameters | Wave 1 | Wave 2 | Wave 3 | Wave 4 |
| --- | --- | --- | --- | --- |
| $\Lambda$ | $1.00 \times 10^4$ | $6.66 \times 10^6$ | $1.59 \times 10^7$ | $1.70 \times 10^7$ |
| $\beta$ | $3.33 \times 10^{-1}$ | $2.49 \times 10^{-1}$ | $1.90 \times 10^{-1}$ | $2.1 \times 10^{-1}$ |
| $\kappa$ | $3.00 \times 10^{-2}$ | $3.45 \times 10^{-2}$ | $4.90 \times 10^{-2}$ | $1.60 \times 10^{-3}$ |
| $\gamma_a$ | $7.00 \times 10^{-2}$ | $1.33 \times 10^{-4}$ | $1.28 \times 10^{-3}$ | $3.90 \times 10^{-2}$ |
| $\gamma_s$ | $3.33 \times 10^{-2}$ | $4.42 \times 10^{-2}$ | $5.10 \times 10^{-1}$ | $2.70 \times 10^{-1}$ |
| $\alpha$ | $1.43 \times 10^{-1}$ | $1.70 \times 10^{-2}$ | $9.00 \times 10^{-3}$ | $2.10 \times 10^{-3}$ |
| $\psi$ | $1.43 \times 10^{-1}$ | $2.45 \times 10^{-1}$ | $9.90 \times 10^{-3}$ | $4.50 \times 10^{-3}$ |
| $\eta$ | $1.60 \times 10^{-2}$ | $4.58 \times 10^{-1}$ | $1.07 \times 10^{-1}$ | $8.50 \times 10^{-2}$ |

The first four plots in Figure 2 can be combined to give the last figure, that depicts the piecewise fitting of the model to the data. What is critical is the initialization of the initial conditions for each wave.

**Figure 2.**
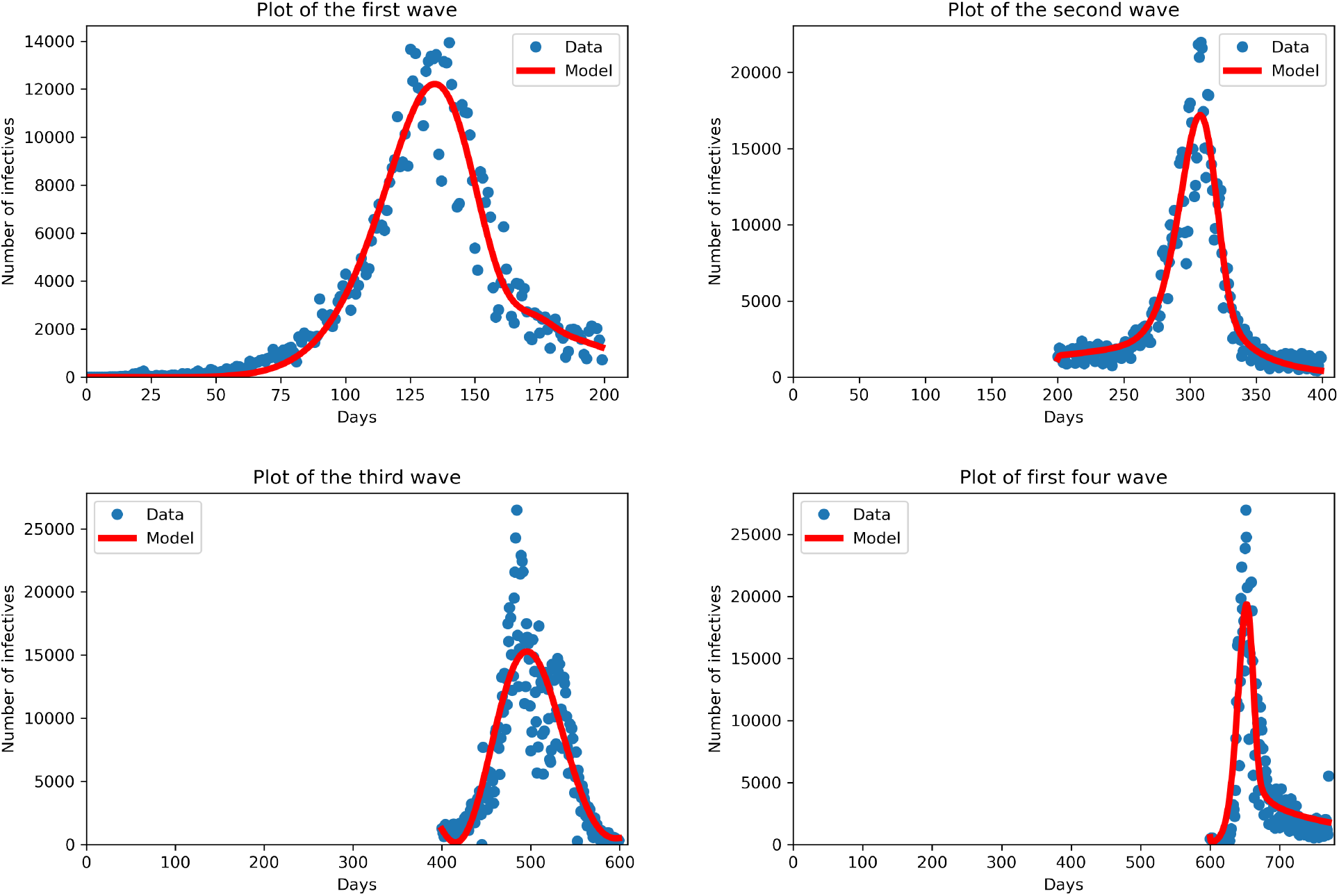
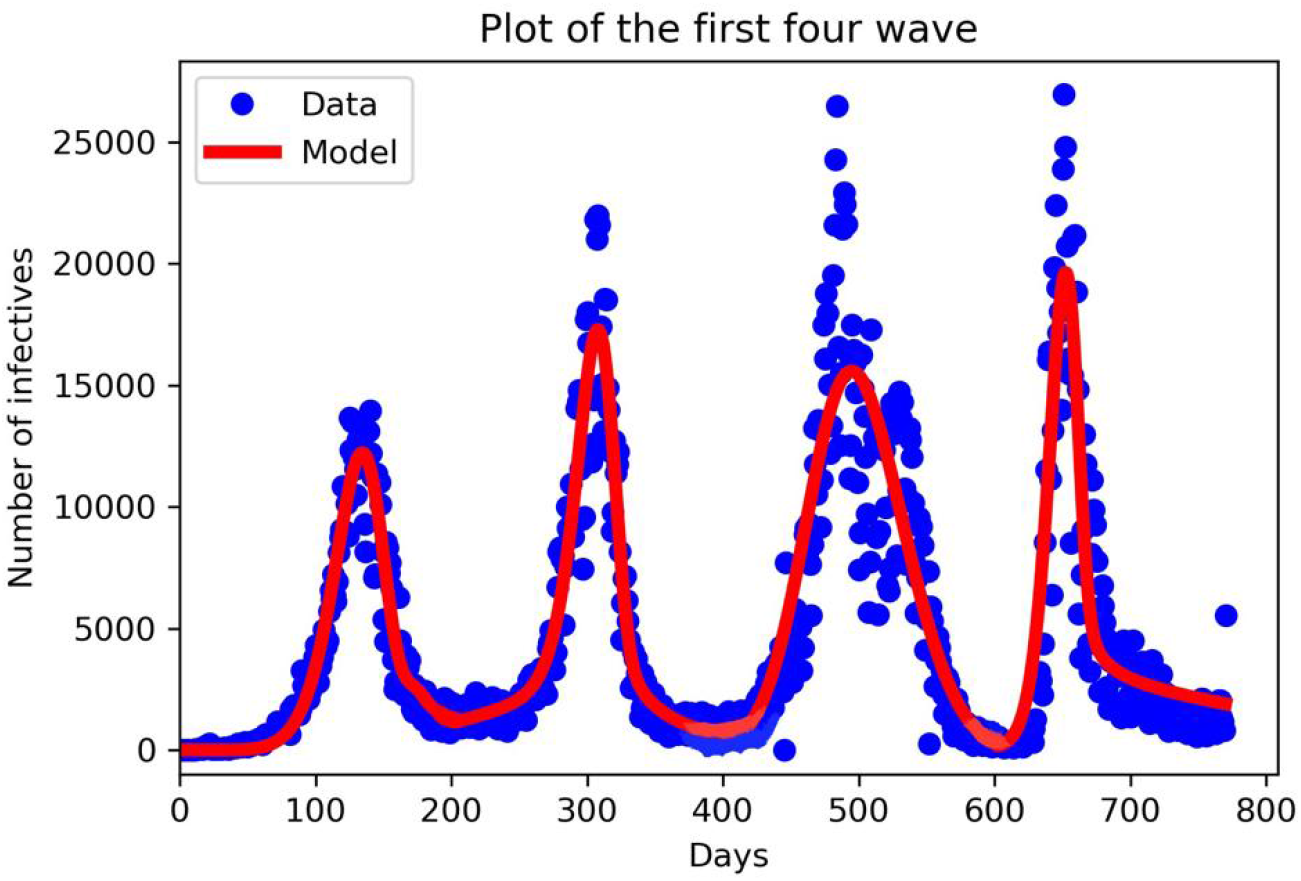
New cases for South Africa for the first four waves. The first wave from 27^*th*^ March to 11^*th*^ September, 2020, the second wave from 11^*th*^ September 2020 to 10^*th*^ April, 2021, the third wave from 11^*th*^ April, to 29^*th*^ October 2021 and the fourth wave ran from 30^*th*^ October 2021 to 19^*th*^ April, 2022. The blue dots denote the new cases from the data and the red lines denote our mathematical model prediction. We can see that in each wave our model constructed fitted well for the given parameters in Table 3.

We conceive that the number of infectives decrease with increasing social distancing over time. In Figure 3, we varying the efficacy of the social distance parameter, when it is 10%, 50% and 90%. Our result shows that, when the efficacy is between 10% − 50%, it increase the disease infection rate since individuals does not observe the rules to social distancing and COVID-19 transmission mode as seen in Figure 3. The disease persists after a long oscillating transient the infection approaches a positive *ω*−periodic solution. When the efficacy is 90% it reduces the infection rate, there reducing the transmission rate too.

**Figure 3.**
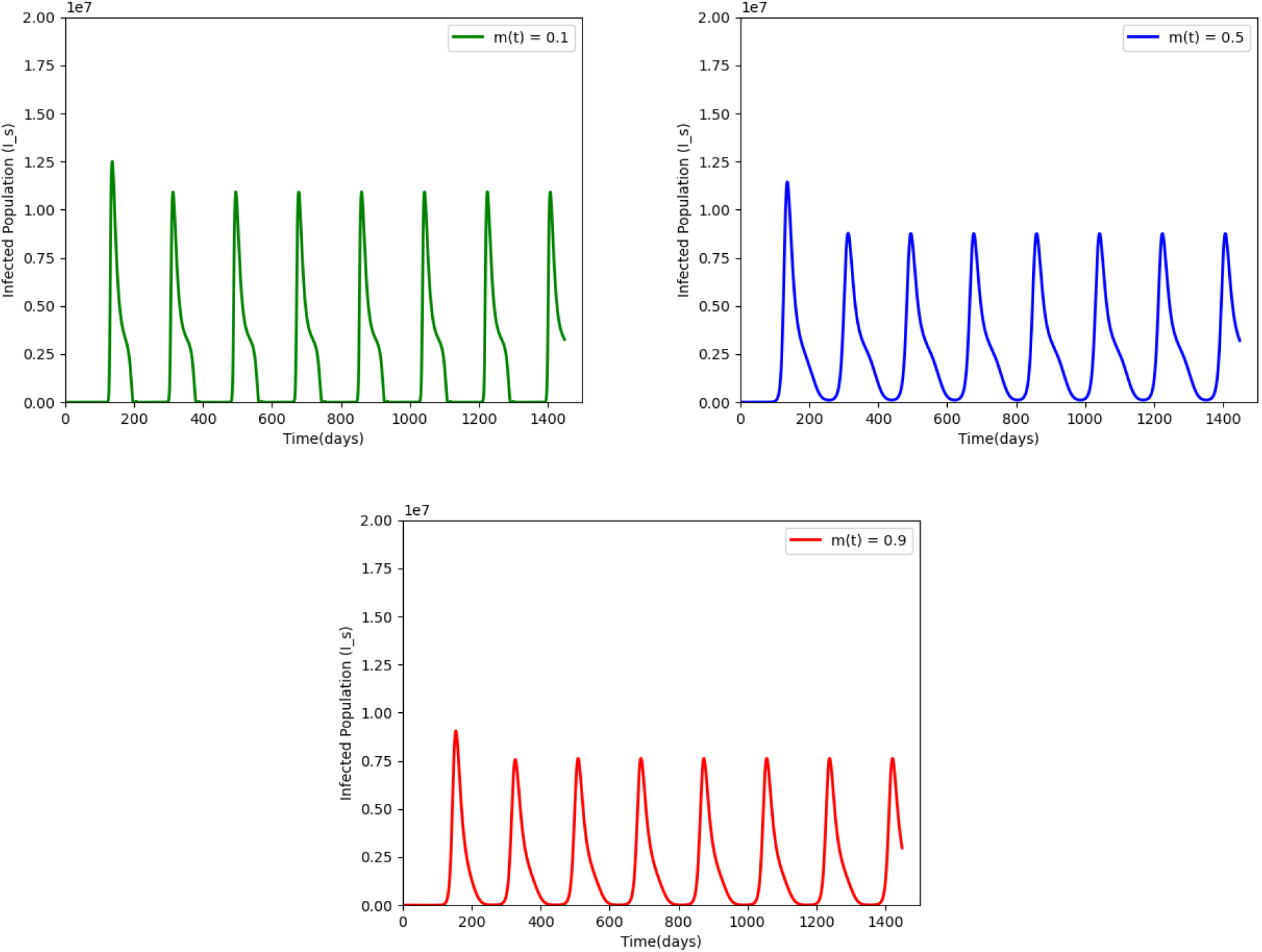
An infection curve of symptomatic individual with social distance parameter with efficacy of 10%, 50% and 90% respectively, in model 2.1, parameters are taken from Table 2, the disease persist and a periodic solution with *ω* = 182.5 days forms after a long transient

## 5. Discussion & conclusion

We proposed a mathematical model to investigate the coronavirus pandemic in South Africa. We included two unique features in our model: the incorporation of social distance parameter in the disease transmission dynamics, and considering it to be periodic due to climate changes or festive seasons experienced in South Africa. We conducted a detailed analysis of this model, and applied it to study the South Africa epidemic using reported data from [26]. Our equilibrium analysis of this model shows that the disease dynamics exhibit a threshold at *R*_0_ = 1. We have established the global asymptotic stability of the disease free equilibrium (*E*^0^) when *R*_0_ < 1, which implies that the disease dies out, and the global asymptotic stability of the endemic equilibrium (*E*^1^) when *R*_0_ > 1, there is at least one positive periodic solution, that is, the disease will spread.

Our numerical simulations results demonstrate the application of our model to the COVID-19 in South Africa and the model fits to the reported data very well. The numerical simulations also suggest that, when raising 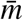 and 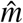 respectively, while keeping the other parameters fixed in the transmission rate increases the infection curve. Also increasing the social distance parameter on a scale of (0 − 1) decreases the infection curve. This means that, climate or festive fluctuations should be taken into consideration in designing policies aimed at COVID-19 control and management.

It is widely speculated that COVID-19 would persist and become endemic. From our analysis and numerical simulations, performed over a time interval of 4 years, the disease was still striving and persisting, supporting this speculation. Our results imply that we should be prepared to fight the COVID-19 for long term than this current endemic wave, in order to reduce the endemic burden and potentially eradicate the disease. We can achieve this when vaccination becomes a requirement.

The model presented in this paper can be improved by incorporating the environmental reservoir into the disease transmission dynamics. When infected individuals cough or sneeze, they spread the virus to the environment through their respiratory droplets which infect other susceptible people with close contact of the same area. The current vaccination drive has the potential of changing the seasonal patterns. However like influenza, a seasonal vaccination program may be a likely scenario in the case of COVID-19 if it continues to be seasonal. This has thepotential of lowering the peaks of the outbreaks and reducing mortality due to the disease.

## Data Availability

All data produced in the present work are contained in the manuscript

## Acknowledgments

The authors are grateful for the support of the Organization for Women in Science for the Developing World (OWSD) and Department of Mathematics and Applied Mathematics, University of Johannesburg for the production of this paper.

